# Performance of early warning signals for disease re-emergence: a case study on COVID-19 data

**DOI:** 10.1101/2021.03.30.21254631

**Authors:** Daniele Proverbio, Françoise Kemp, Stefano Magni, Jorge Gonçalves

## Abstract

Developing tools for rapid and early detection of disease re-emergence is important to perform science-based risk assessment of epidemic threats. In the past few years, several early warning signals (EWS) from complex systems theory have been introduced to detect impending critical transitions and extend the set of indicators. However, it is still debated whether they are generically applicable or potentially sensitive to some dynamical characteristics such as system noise and rates of approach to critical parameter values. Moreover, testing on empirical data has, so far, been limited. Hence, verifying EWS performance remains a challenge. In this study, we tackle this question by analyzing the performance of common EWS, such as increasing variance and autocorrelation, in detecting the emergence of COVID-19 outbreaks in various countries. We illustrate that EWS are successful in detecting disease emergence if some basic assumptions are satisfied: a slow forcing through the transitions and not-fat-tailed noise. In uncertain cases, noise properties or commensurable time scales may obscure the expected early warning signals. Overall, our results suggest that EWS can be useful for active monitoring of epidemic dynamics, but that their performance is sensitive to certain features of the underlying dynamics. Our findings thus pave a connection between theoretical and empirical studies, constituting a further step towards the application of EWS indicators for informing public health policies.

**Author summary:** To extend the toolkit of alerting indicators against the emergence of infectious diseases, recent studies have suggested the use of generic early warning signals (EWS) from the theory of dynamical systems. Although extensively investigated theoretically, their empirical performance has still not been fully assessed. We contribute to it by considering the emergence of subsequent waves of COVID-19 in several countries. We show that, if some basic assumptions are met, EWS could be useful against new outbreaks, but they fail to detect rapid or noisy shifts in epidemic dynamics. Hence, we discuss the potentials and limitations of such indicators, depending on country-specific dynamical characteristics and on data collection strategies.

## 1 Introduction

Epidemics such as the current COVID-19 pandemic pose important and long-lasting threats to human societies [1]. Hence, developing tools for rapid and early detection of disease emergence is important to perform science-based risk assessment [2]. In principle, detailed mechanistic understanding could help formulate predictive models. However, combinations of non-linearity, noise and a lack of curated datasets for validation hamper the development of mechanistic models. Therefore, numerous recent studies have suggested using different methods, agnostic of detailed mechanistic models, that could detect shifts in epidemic dynamics [3, 4, 5, 6, 7]. These methods are based on the theory of critical transitions in dynamical systems [8] and require the calculation of statistical early warning signals (EWS) from observed data. However, the applicability of such early warning signals is still debated, as it might depend on the interplay of modelling predictions and empirical observed dynamics.

Critical transitions encompass a broad class of complex phenomena characterized by sudden shifts in the system dynamics. Key mechanisms for deterministic shifts are dynamical bifurcations [9], i.e. qualitative changes of equilibria due to leading eigenvalues crossing a threshold value. In epidemiology, the leading parameter is the reproduction number *R*, the average number of secondary infections from a single contagious case in a susceptible population [10, 11]. Re-emergence of infectious diseases thus involves a transmission system that is pushed over the critical point *R* = 1 through a transcritical bifurcation [7]. As a consequence, it may in principle be possible to apply results from the theory of critical transitions to detect impending epidemic re-emergence. In particular, proposed early warning signals (EWS) are summary statistics indicators that might change in a predictable way when approaching the critical threshold. Common EWS are increasing variance and autocorrelation, which have been suggested to be generically applicable to detect impending regime shifts in different systems [12, 13]. If this would be the case, the consequences would be far-reaching, possibly expanding the set of epidemic indicators, using complexity theory. Several theoretical and computational studies already investigated EWS performance on abstract epidemiological models [7, 4, 14, 15, 16], but so far only a few testings on empirical data have been performed [17]. Performing more tests is a necessary next step towards the application of EWS in routine surveillance procedures.

In this study, we aim at testing the performance of EWS in detecting the re-emergence of observed epidemics, and at explaining the observed performances based on the correspondence of modelling assumption and dynamic features observed in the data. Specifically, we are not screening all possible EWS on all possible empirical data [18, 19]; on the opposite, we are testing whether some EWS work when they are expected to, what happens in other cases, and why. In fact, theoretical predictions like early warning signals should ideally be tested in controlled experiments [20], but these are often not feasible in complex phenomena like epidemics. Instead, the present work considers curated observational data from many outbreaks of the same disease, following the strategy of “natural experiments” [21]: first, constructing a dataset that includes relevant time series data; second, accounting for possible confounders, i.e. dynamical characteristics that might alter the expected signals; third, evaluating the performance of EWS and interpreting it in light of previous theoretical results.

To this end, we use worldwide data from the current COVID-19 epidemic. The COVID-19 disease, caused by severe acute respiratory syndrome coronavirus 2 (SARS-CoV-2) [22], rapidly diffused in the whole world during 2020 and 2021. After a first outbreak, countries worldwide managed to curb the local infection curves with combinations of non-pharmaceutical interventions, which tuned the effective time-dependent reproduction number *R*(*t*) to values below 1 [23]. Later during the year 2020, many countries experienced epidemic re-emergences (often called “second waves”) associated to *R*(*t*) re-crossing 1 from below [24, 25]. The unprecedented diffusion of the virus, as well as the various degrees of intervention strengths, provide abundant epidemic data to construct the test set. Known dynamical features associated to modelling assumptions, such as noise and rate of evolution of *R*(*t*) [26], are then accounted for by analysing the time series of each country. These features allow to interpret the trends observed in empirically derived EWS and their performance in different contexts.

The paper is organised as follows. First, we recall theoretical results from literature, to allow the subsequent comparison with expected EWS behavior. Then, we describe how the test set was constructed and analysed. After that, we study the behavior and the performance of EWS from empirical data and their dependence to dynamical characteristics associated with modelling assumptions. Finally, we discuss the current findings, their limitations and their implications for future studies.

## 2 Methods and Mathematical Theory

### 2.1 Mathematical theory and derivation of EWS

The scope of this article is to test theoretical predictions in light of their assumptions. Hence, we provide a brief review of the theoretical basis of critical transitions in epidemic dynamics, as well as on the derivation and assumptions that underlie their associated early warning signals. This way, we highlight the theoretical results to be tested as well as their supporting hypothesis, which will be central for this study. Further details can be found in the supporting information material S1 as well as in the references provided.

It is often recognised that epidemics can be described as complex systems, whose macro-scale dynamics evolves out of equilibrium [3, 7, 13]. In different complex systems, sudden dynamical changes can happen when the system is pushed over a critical point through a bifurcation [27]. Critical transitions observed in complex systems are often associated with such bifurcations [28]. Recent studies have shown that the trend of certain statistical indicators may signal the approach to a critical transition in slowly forced dynamical systems [9, 29]. In general, a slowly forced system with variables **x** and control parameter *q* is [30]:

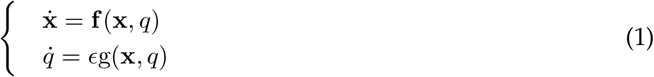

with 0 *< E* « 1. Typical models of epidemic dynamics, such as SIR-like models [23], can be expressed as a slowly forced system like Eq. 1 when the control parameter R slowly approaches its critical value 1.

In the presence of small random fluctuations, the approach to the critical point is often associated with predictable trends of several statistical indicators of variability, which have been proposed as early warning signals of impending transitions [31, 32]. The reason is that noise can push the system state around the deterministic trend of Eq. 1; at the same time, its statistical properties might change as the system approaches the transition and could be used to detect it [30]. If the noise is relatively small with respect to the deterministic trend and normally distributed, the trend of the most common summary statistics (variance, autocorrelation, skewness, coefficient of variation), computed on detrended residuals, is expected to increase next to the transition, thus providing an early warning signal [13, 33]. If this signal can be observed prior to the transition, it would constitute an early warning.

Several early warning signals from the critical transitions theory have been predicted to apply on epidemiological models. A particularly relevant theoretical result is from [7]. The authors consider an extended SIR model:

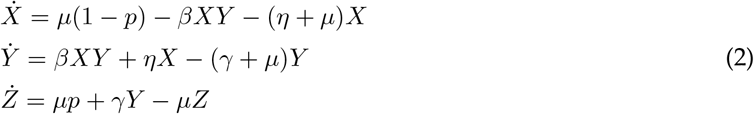

with variable *X* for susceptible, *Y* for infectious, *Z* for removed. *β* represents the infection rate and *γ* the removal rate [34]; *µ* describes the flux of people across country boundaries; *η* is an influx of infected cases that could trigger a new infection; *p* represents a protection rate for the susceptible population, either by noon-pharmaceutical interventions or by vaccination [35]. In this case, the reproduction number is [7]:

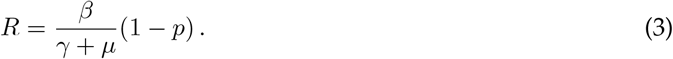

There is a transcritical bifurcation on the (*Y, p*) diagram when *R* reaches its critical value *R* = 1 for *p*^*\**^ = 1 − (*γ* + *µ*)*/β*. When considering the stochastic version of Eq. 2, it is possible to analyse its fluctuations next to the transition and extrapolate the early warning signals from the associated summary statistic indicators. The kind of noise (additive or multiplicative) that is considered constitutes a modelling assumption. How the most common indicators — variance and lag-1 autocorrelation — are derived (from [7]) is reported in the supporting information, along with their predicted evolution next to the critical transition when the slow-fast assumption is satisfied, or not (Fig. S1). Their increasing trends prior to the transition provide the predicted early warning signal.

Further computational results, which also consider additional indicators, suggest that the increases in variance are the best performing indicators of re-emergence, in terms of signal-to-noise ratio and of detection performance [3, 4]. However, as noticed in follow-up studies [30, 36, 37], their performance is linked to whether their modelling assumptions are satisfied. If it is the case, such indicators perform well; but what happens in other contexts is still less clear.

We here recall the main modelling assumptions underlying the prediction of EWS, as well as their relevance for their performance. (1) critical transitions are local phenomena. Hence, EWS are not global measures, but are expected to work in the vicinity of the regime shift. (2) it should be possible to express the epidemic dynamics in terms of a fast-slow system like Eq. 1. When approximating *R* approaching to 1 as a linear trend, the modelling assumption Eq. 1 is satisfied if the regression coefficient (the slope of the linear trend) is small. Otherwise, literature results suggest that the expected patterns will be either distorted or will not occur [3, 7]. (3) the closer random fluctuations are to be additive noise, the more robust the performance of EWS is. If there are deviations from white noise, EWS trends can be modified or disrupted. For instance, decreasing variance was observed in case of non-white multiplicative noise [37]. (4) In case of combinations of non-white noise and of non-fast-slow description, one might observe bifurcation delays, i.e. changes of the system state (and of its indicators) that lag behind the theoretical bifurcation. This would translate in a warning signal that emerges much time later than the epidemic reemergence. (5) If the transition is triggered by large random fluctuations — the so called noise-induced transitions [9] — no EWS is expected to be observed [38].

### 2.2 Data collection and curation

This paper studies the re-emergence of infectious diseases in a number of observations from all over the world. Our aim is to verify whether EWS work when they are expected to and explain why they might malfunction otherwise, rather than perform an observational study over all COVID-19 re-emergencies (for such a study, refer to [18]). Consequently, to construct the dataset, we considered data from countries that faced a re-emergence of positive COVID-19 cases between beginning of March (starting of wide viral diffusion) to mid-September 2020. We did not consider further data points as many countries began issuing new social measures that rapidly impacted the epidemic trends. These would hinder the careful analysis of confounders.

When possible, we use prevalence data, i.e. active cases, in accordance to what is modeled by SIR-like models and to what was suggested in literature [14]. Active cases from Luxembourg are retrieved from the government website (COVID19.public.lu/fr/graph). When not directly available, active cases *A* are estimated, following [39], by the proxy:

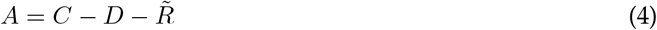

where *C* indicates the cumulative positive cases, *D* the number of registered deaths and 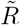 the number of recovered patients. Country data are obtained from the John Hopkins University collection [40] and the European Centre for Disease Prevention and Control database (https://www.ecdc.europa.eu/en/COVID-19/data). We also use Italian data from the Veneto region, as an example of regional data with an identifiable second wave during the considered time interval. Veneto time series are retrieved from the Github repository of the Italian “Dipartimento della Protezione Civile - Emer- genza Coronavirus” (https://github.com/pcm-dpc/COVID-19). All databases are accessed up to 15/09/2020.

To best curate the database, an initial screening on data quality is performed. We reject time series with very few active cases, as mostly driven by purely stochastic processes. We also discard time series for which the share of positive cases over performed tests is *>* 5% next to the transition, as WHO guidelines suggest possible undertesting (we refer to WHO reports such as https://bit.ly/3dARcy1). Information about the share of positive tests is obtained from the OurWorldInData curated dashboard [41]. As EWS from critical transitions are based on mean-field homogeneous SIR-like models, we do not consider whole countries with clear spatial heterogeneity like Italy [42], but we instead use regional data if available. Finally, we discard some public time series that behave clearly differently from the common “bell-shaped” epidemiological curve [39] (see electronic supporting information Fig. S2).

### 2.3 Analysis of dynamical features

To identify the transition *a posteriori* and get a “ground truth” date of re-emergence, we use a data-driven estimation of the time-dependent *R*(*t*). Similar to [4, 43, 44], *R*(*t*) is estimated with a Bayesian Markov Chain Monte Carlo method (see also electronic supporting information Sec. S4).

Then, we employ the posterior Bayesian estimate 𝒫 (*R*(*t*) *>* 1) to assess the probability that the control parameter is greater than 1. Since *R*(*t*) *>* 1 is associated with an exponential increase of infectious cases after a transcritial bifurcation, 𝒫 (*R*(*t*) *>* 1) can be interpreted as the probability of seeing an epidemic outbreak. Such probability is defined as:

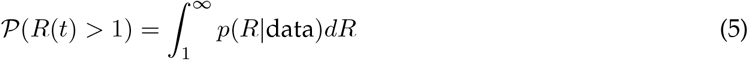

where *p*(*R* | data) is the posterior probability density function obtained with the Bayesian framework. The most likely day *t*_*em*_ in which the transition happened, assumed as our ground truth, corresponds to the maximum of 𝒫 (*R*(*t*) *>* 1) from Eq. 5 (see electronic supporting information Fig. S3).

After calculating the outbreak date, we test the modelling assumptions of normally distributed fluctuations and of slow approach to the critical transition.

To test the additive noise assumption, we analyze the global distribution of stochastic fluctuations, filtered from the time series with a 7-days moving Gaussian kernel as suggested in [45, 46]. The window size reflects typical cycles of data reporting and of COVID-19 fluctuations [47]. The distribution of fluctuations over the complete time series is indicative of the average noise distribution. We computed skewness and kurtosis to measure deviations from Gaussian noise, which is characterized by skewness=0 and kurtosis=3 [48].

To test the assumption of slow approach to the transition, we measure the rate of approach of the control parameter to its critical value. For this, we compute the time-dependent *R*(*t*) like above and, consistently with the fast-slow system description Eq. 1, we fit a linear function:

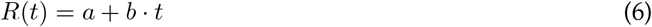

in the interval 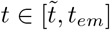 Here, *t*_*em*_ corresponds to the day associated with novel disease emergence as explained above; 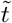 is the day associated with the minimum of *R*(*t*) after the first wave. The regression coefficient *b ± σ*_*b*_ measures the ramping speed of the control parameter (along with its uncertainty). As an indication, *R*(*t*) is said to be “slowly evolving” if it goes from its minimum value to 1 in a period of time that is much longer than the COVID-19 serial interval (around 4 days [49]), which is a proxy of the disease duration time scale. For the fitting, we use the *scipy* Python library. Refer to electronic supporting information Sec. S5 for details.

### 2.4 Estimation of EWS

Estimation of early warning signals from time series data is performed following standard methods from literature [26].

First, we detrend the time series to obtain a moving average, representative of the deterministic trend. The “residuals” or detrended fluctuations are obtained by subtracting the moving average from the original time series. To investigate possible effects of detrending approaches — as discussed in previous theoretical studies [13, 29, 45] — we use and compare three detrending methods: a uniform moving mean, a Gaussian kernel, and ARIMA models [50]. The ARIMA models are specifically tuned for each country, see electronic supplementary material.

Then, we compute the statistical indicators associated to each point with a backward sliding window, i.e. one where the associated time point is the rightmost one. This way, all estimates are agnostic of future values. All EWS indicators are estimated on the detrended time series. We initially calculate the variance, which is suggested to be the most robust indicator for epidemic re-emergence [3, 4, 7, 14] as:

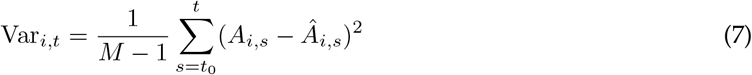

for any time point *i* with active cases *A*, over a sliding window with size *t* − *t*_0_ including *M* time points. Â is the moving average. We also estimate other common statistics such as lag-1 autocorrelation AC(1), coefficient of variation (CV) and skewness, which are constructed similarly to the variance over the same sliding window. The sampling frequency of COVID-19 data is not sufficient to allow estimation of the power spectrum reddening [51] or of the sample entropy [3]. All indicators are estimated with their corresponding Matlab functions. Note that the estimation of *𝒫*(*R*(*t*) *>* 1) is done a posteriori, that is, once we know the complete time series. Instead, the early warning signals are calculated a priori, without knowing in principle if a transition is approaching.

### 2.5 Quantification of EWS trends and Receiver Operator Characteristics analysis

Recent studies [14, 26, 36] suggest to quantify the expected increasing trend of EWS next to the transition with the Kendall’s *τ* coefficient of monotonicity. The Kendall’s *τ* score is defined as [52]:

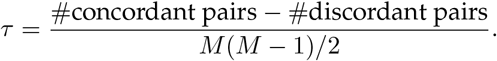

*M* is the number of considered time points. Two generic points (*t*_1_, *x*_1_) and (*t*_2_, *x*_2_) are said to be a concordant pair if, for *t*_1_ *< t*_2_, *x*_1_ *< x*_2_, and a discordant pair otherwise. A constant trend is expected to have *τ* = 0. We compare this value with the *τ* scores calculated on time series with identified transitions. To go beyond simple visual inspection, we quantify the detection power of each statistical indicator using its time-changing trend, classifying data as either belonging to the second wave or not. We calculate each EWS on a moving window (its size is discussed later in the text) for each detrended time series, and compare the Kendall’s *τ* score calculated over a period −30 *< t <* 5 days around the transition. *t >* −30 is chosen to avoid significant overlaps with the first epidemic wave, *t <* 5 to account for possible small bifurcation delays [13].

We use Receiver Operator Characteristics (ROC) analysis to classify each time point as either before or after re-emergence. We compare each statistical indicator’s ability to correctly distinguish which Kendall’s *τ* scores belong to those from before or after re-emergence. To do so, we compare various values from 0 *< τ <* 1 to that next to the transition, for each country. To increase the number of transitions to test and estimate the mean detection performance of an indicator, we average over all countries in a test set. The ROC analysis returns the Receiver Operator Characteristics (ROC) curve, a parametric plot of the sensitivity and specificity of a classification method as a function of the detection threshold [4, 53]. The overall detection performance of each EWS is quantified by the area under the ROC curve (AUC). A value AUC= 0.5 means that the statistics detection performance is as good in classifying as randomly guessing. A good indicator should have AUC close to 1, which informs that it is possible to identify the transition by the increasing trend of the indicator. An AUC close to 0 indicates good classification, although resulting from a decreasing indicator that does not correspond to the predetermined theoretical prediction.

## 3 Results

### 3.1 Analysis of country-wise dynamical characteristics associated to the spread of COVID-19

Tab. 1 reports the list of countries that satisfy the curation requirements discussed in Materials and Methods and are thus included in the analysed datased. Tab. 1 also reports the dates of re-emergence, identified by the analysis of *R*(*t*). Fig. 1a shows an example of time series of active cases for Luxembourg, from March to mid-September 2020, with the date of estimated re-emergence (dashed line).

**Table 1:** Selected countries for the dataset, abbreviations and date of second epidemic insurgence. Refer to “Data Collection and Curation” for how the date marking the second wave is obtained.

| Country | Abbr. | Date |
| --- | --- | --- |
| State of Victoria (Australia) | AUS | 27/06/2020 |
| Austria | AUT | 01/07/2020 |
| Denmark | DNK | 03/08/2020 |
| Israel | ISR | 01/06/2020 |
| Japan | JPN | 28/06/2020 |
| Korea, South | KOR | 13/08/2020 |
| Luxembourg | LUX | 29/06/2020 |
| Nepal | NPL | 29/07/2020 |
| Singapore | SGP | 25/07/2020 |
| Veneto (Italy) | VEN | 29/07/2020 |

**Fig. 1.**
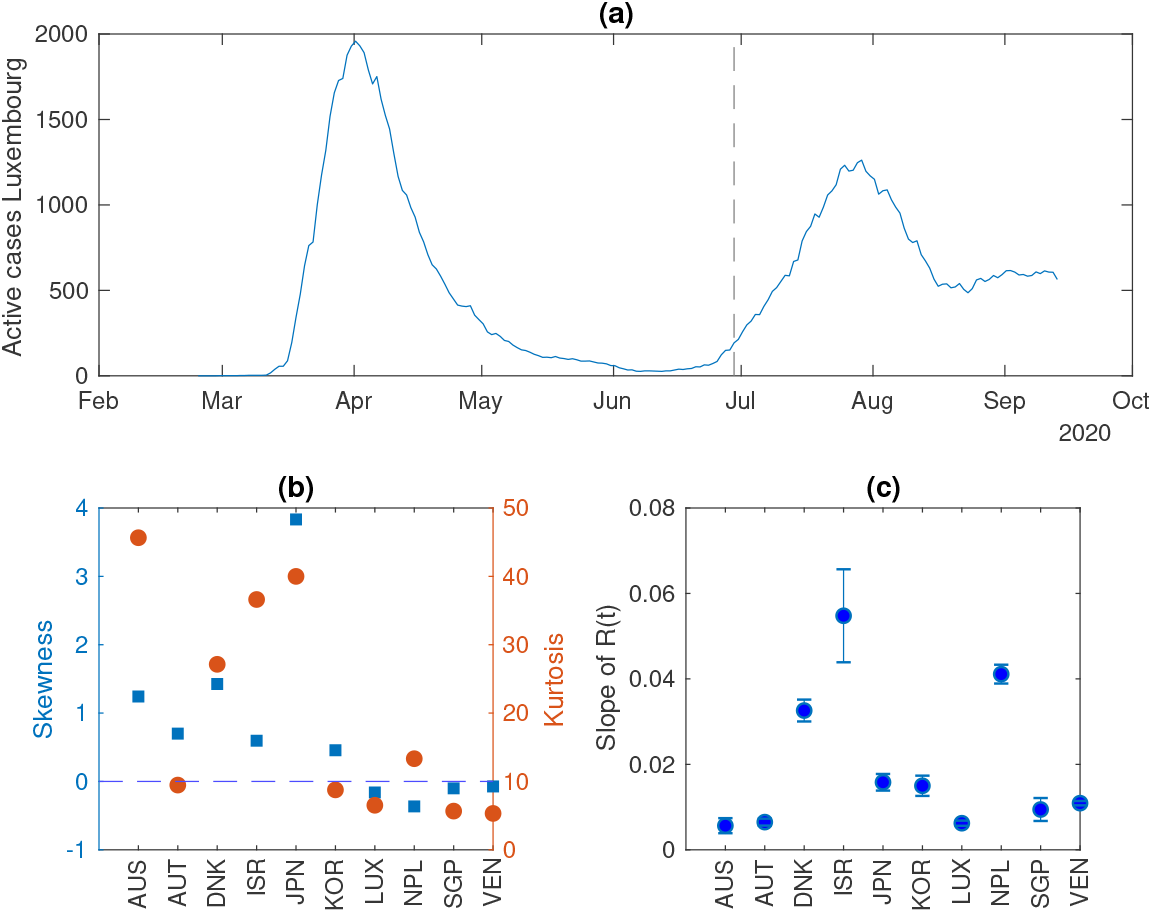
Analysis of the dynamical characteristics of the countries included in the test set 𝒴. a) An example of an epidemiological curve of active cases. The dashed line indicates the transition, measured by *R >* 1. The latter is objectively identified by the date at which the probability of *R*(*t*) to be greater than 1 is at its maximum (see “Analysis of dynamical features”); b) Measures of the distribution of data fluctuations. Skewness *µ* indicates the symmetry of the distribution, whereas kurtosis *γ* indicates the relevance of its peak with respect to the tails. Large deviations from *µ* = 0 (dashed line) and *γ* = 3 are associated with non-normal distributions. c) The regression coefficient of *R*(*t*) and its associated uncertainty, as obtained from the linear fit Eq. 6.

The time series of the considered countries show different dynamical features. Fig. 1b shows various ranges of noise distribution, measured by skewness and kurtosis. Austria, Luxembourg, Nepal, Singapore and Veneto display noise distributions that are close to Gaussian, while noise in the other countries is further away from white than in the previously mentioned ones. This could be associated to social dynamics or imperfect data reporting [54].

The rate of approach of *R*(*t*) to its critical value also differs, as indicated in Fig. 1c by the regression coefficient of a linear fit for *R*(*t*) (*cf*. Eq. 6). State of Victoria, Austria, Luxembourg, Singapore and Veneto display a slow approach to the critical value and can thus be better suited to be appropriately described as slow-fast systems like Eq. 1. Japan and South Korea show intermediate values, while other countries - Denmark, Israel and Nepal - have a faster evolution of the control parameter, which does not satisfy the assumption of slow evolution.

By this analysis, we notice that Luxembourg satisfies the modelling assumptions and is the closest to being a “controlled experiment” according to the criteria described in the section about deriving EWS. In fact, the country is small, homogeneous population-wide interventions were in place, and a Large Scale Testing (LST) strategy was implemented to best monitor the virus diffusion in the country [55]. This country-wide testing strategy reached more than 70.000 tests per week over a population of about 600.000, thus allowing extensive and frequent monitoring.

Other countries (State of Victoria, Austria, South Korea, Singapore, Veneto) satisfy some of the modelling assumptions. We thus include them in a test set 𝒴, used to further assess the performance of EWS. In fact, they either display a similar rate of approach to the transition as Luxembourg (within the error bars), or a similar noise distribution.

The remaining countries (Denmark, Israel, Japan, Nepal) do not satisfy either of the modelling assumptions and are grouped in a set 𝒩. This is used to interpret the performance of EWS in settings that are not properly described by theoretical models and represent possible limitations of the predetermined predictions.

### 3.2 Local trends on controlled data and impact of detrending methods

We first focus on Luxembourg, that displays the best data, to test the theoretical predictions about the local behavior of common EWS. Summary statistical indicators are estimated from the detrended fluctuations (residuals) around prevalence data as per standard methods [26].

We first investigate the effect of the detrending method in generating residuals. To do so, we compare the fluctuations around the deterministic trend obtained with a Gaussian kernel smoothing [45], a moving average filtering [13] and an ARIMA(2,1,3) model [50]. Fig. 2 shows the time evolution of the residuals obtained with the three methods, as well as their mutual correlations. As quantified by the Pearson correlation coefficient, the Gaussian and moving average filtering have similar output (correlation coefficient *ρ* = 0.95); hence, the Gaussian kernel smoothing is used in the rest of the analysis. However, the ARIMA method returns residuals that are less correlated wit the previous ones (*ρ* = 0.23), whose effect on EWS needs further investigation.

**Fig. 2.**
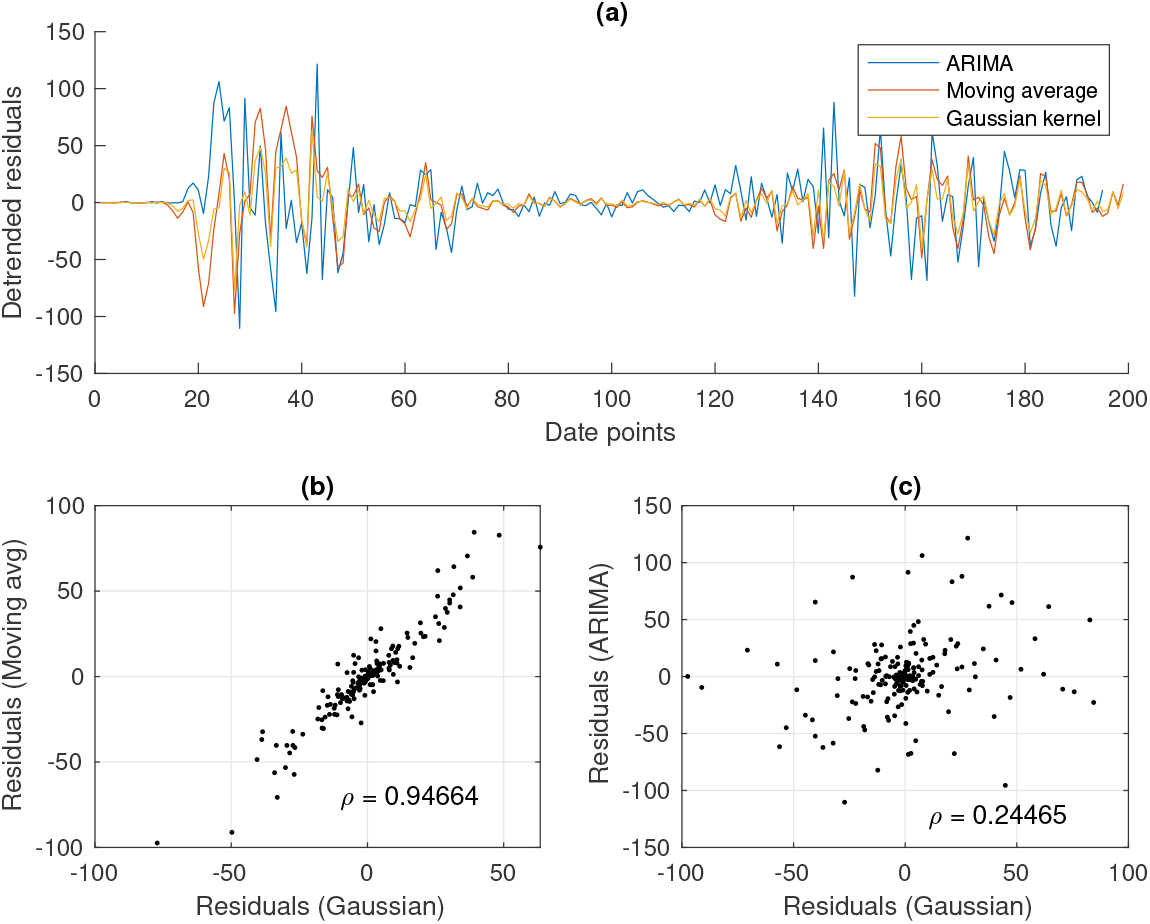
Analysis of the residuals from the smoothing methods. a) The detrended fluctuations time series. b) Correlation between residuals obtained from Gaussian or moving average filtering. c) Correlation between residuals obtained from Gaussian or ARIMA filtering.

Then, we study the behavior of the variance (theoretically, the most robust EWS [4]) next to the transition point, identified as the day when the estimated *R*(*t*) crosses 1 (dashed line in Fig. 3). The increase in variance prior to the transition, as expected from theoretical studies, is evident in Fig. 2, irrespective of the moving window size and on the detrending method. Although the lead time is slightly advanced for shorter window sizes, the corresponding Kendall’s *τ* measure of monotonous increase is similar for both methods and all window sizes (*cf*. values reported in Fig. 3). In general, a large window size produces a visually reduced absolute increase, whereas a small window size is associated with less smoothed curves. From here on, we will use a window of 14 days as a reasonable trade-off, collecting enough data to be robust without being over-dependent on past history. The ARIMA residuals produce a visually clearer increase in variance, but the Kendall’s *τ* quantifies an analogous trend (even slightly lower). Hence, for the incoming quantitative analysis on the EWS performance, we concentrate on data obtained from the Gaussian kernel filtering.

**Fig. 3.**
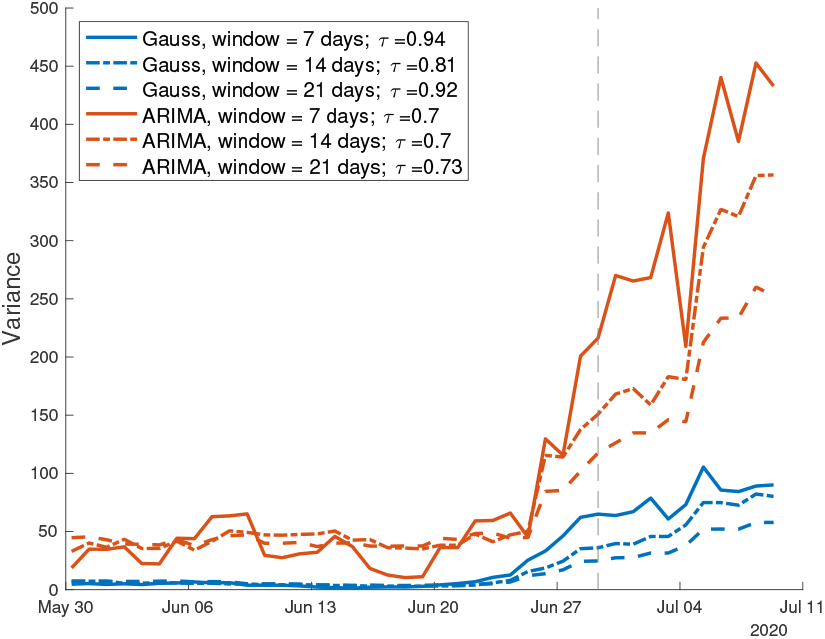
Analysis of the variance in the Luxembourg controlled setting. Its increase is evident prior to the transition (dashed line). The increasing trend is little sensitive to the sliding window size, as displayed by the three curves and by *τ* values reported in the text. The variance is computed over the residuals from Gaussian filtering and ARIMA detrending. The increasing trend during the considered time window is quantified by the associated *τ* values.

These findings confirm that, in a controlled setting that satisfy the modelling assumptions, the theoretical predictions on EWS are valid. Indeed, an increasing trend of the variance in the vicinity of the transition point could serve as early warning to detect the transition to disease emergence.

### 3.3 Global trends of EWS

After confirming the local behavior of the variance in a highly controlled setting, we widen the analysis to the global performance of other EWS, *i*.*e*. far from the bifurcation, and for different countries from the pre-defined dataset, *cf*. Tab. 1. This way, we further test the theoretical predictions and the EWS potential use in more general contexts.

Among the indicators, we estimate lag-1 autocorrelation (AC(1)), skewness and coefficient of variation (CV), which are often proposed as alternatives to the variance. The size of the moving window is set to 14 days as discussed above. To compare the trend of EWS with the approach to the bifurcation, the probability of *R*(*t*) to be greater than one (from Eq. 5) is also calculated and reported.

Fig. 3 shows the results for Luxembourg, Austria, State of Victoria (from the test set 𝒴). In addition, Israel, which does not satisfy the EWS assumptions (*cf*. Fig. 1) is reported to inspect a deviant case. The figure focuses on EWS trends after the first wave, up to more than a month after the second epidemic insurgence. The graphs for other countries from Tab. 1 are reported in electronic supporting information Fig. S5, along with their associated prevalence data and estimated *R*(*t*).

Focusing first on Luxembourg and Austria, the variance follows its theoretically predicted behavior closely (*cf*., e.g., Fig. 9b in [7] and Fig. S1), with a small but visible increase prior to the transition and a subsequent monotonous trend along the second wave. In Austria, though, it still displays some fluctuations after the relaxation of the first wave. The same happens for the coefficient of variation CV; this is expected, as it depends on the variance and on a stable equilibrium in infectious numbers. On the other hand, the lag-1 autocorrelation shows an increasing trend very close to the transition point, but gives possibly spurious signals during the global time series. Finally, the skewness does not display immediately detectable relevant trends, as anticipated by computational studies [14]. This might be related to noise properties, as suggested by [56].

Variance and CV on Australian data, when processed by eye, show a delay in their increasing trend, which begins around the 7^*th*^ of July (here, we consider the expected *small* increase like the one in LUX and AUT). This is likely a footprint of the so-called “bifurcation delay”, which is associated with deviations from Gaussian noise [13, 37] as those reported in Fig. 1. In this case, the delay is of about two weeks, which could impair online detection of epidemic re-emergence.

Finally, Israel provides an interesting case study as it diverges from the theoretical assumptions, see Fig. 1. In fact, its transition to epidemic re-emergence is rapid, and the noise distribution is far from being Gaussian. These characteristics disrupt the EWS trends as predicted by the theory. In fact, the variance remains flat around the transition, CV and skewness slightly decrease, while lag-1 autocorrelation does not display informative patterns. A bifurcation delay is reported more than 20 days after the transition, but it is as abrupt as the exponential increase in infectious data (see also electronic supporting information). This shows that the application of early warning signals indicators on correct contexts is crucial to obtain reliable signals for developing risk assessment analysis.

### 3.4 ROC quantitative analysis of EWS performance

For the online detection of incoming re-emergence, distinguishing between robust increases and spurious fluctuations is crucial to optimise the true positive signals and minimise the false negatives. Hence, a retrospective analysis of time data is often not sufficient and is only useful for offline detection. Hence, we provide a quantitative estimation of EWS performance in robustly detecting the transition. The Kendall’s *τ* score is used to evaluate if a certain indicator corresponds to an increasing or decreasing trend and compare this for different data types [36, 14]. Hence, we evaluate *τ* for each indicator, over the same 14 days window, and we assess which values are associated with a passage through the transition point. The increase in *τ* is reflected in the Receiver Operator Characteristics (ROC) curve and quantified by Area Under the Curve (AUC) scores. Fig. 4 shows the ROC curves for the considered indicators averaged over the countries in 𝒴. Panel (a) reports ROC curves calculated over data detrended with Gaussian filtering; panel (b) focuses on ARIMA detrended data. Tab. 2 reports the corresponding AUC values, for both methods. The variance is the only indicator that consistently performs better than a random classifier, while the lag-1 autocorrelation performs slightly better for a number of null values *τ*_0_ *>* 0.5. This validates aforementioned theoretical results from literature, e.g. [14, 4]. Instead, the skewness does not improve the detection performance. This is probably due to its fluctuations around the 0 value, as noticed in Fig. 3, which is in turn associated with noise distribution of original data. Interestingly, the coefficient of variation is overall the worst performer. We speculate that this is due to its sensitivity to data fluctuations, which are often non negligible even in countries that belong to the test set 𝒴 (*cf*. Fig. 1). We acknowledge that our findings are sensitive to the estimated time of emergence, which also complicates the estimation of the lead time. Currently, the best lead time is of 5 days for Luxembourg, a setting that is close to the analytical assumptions.

**Fig. 4.**
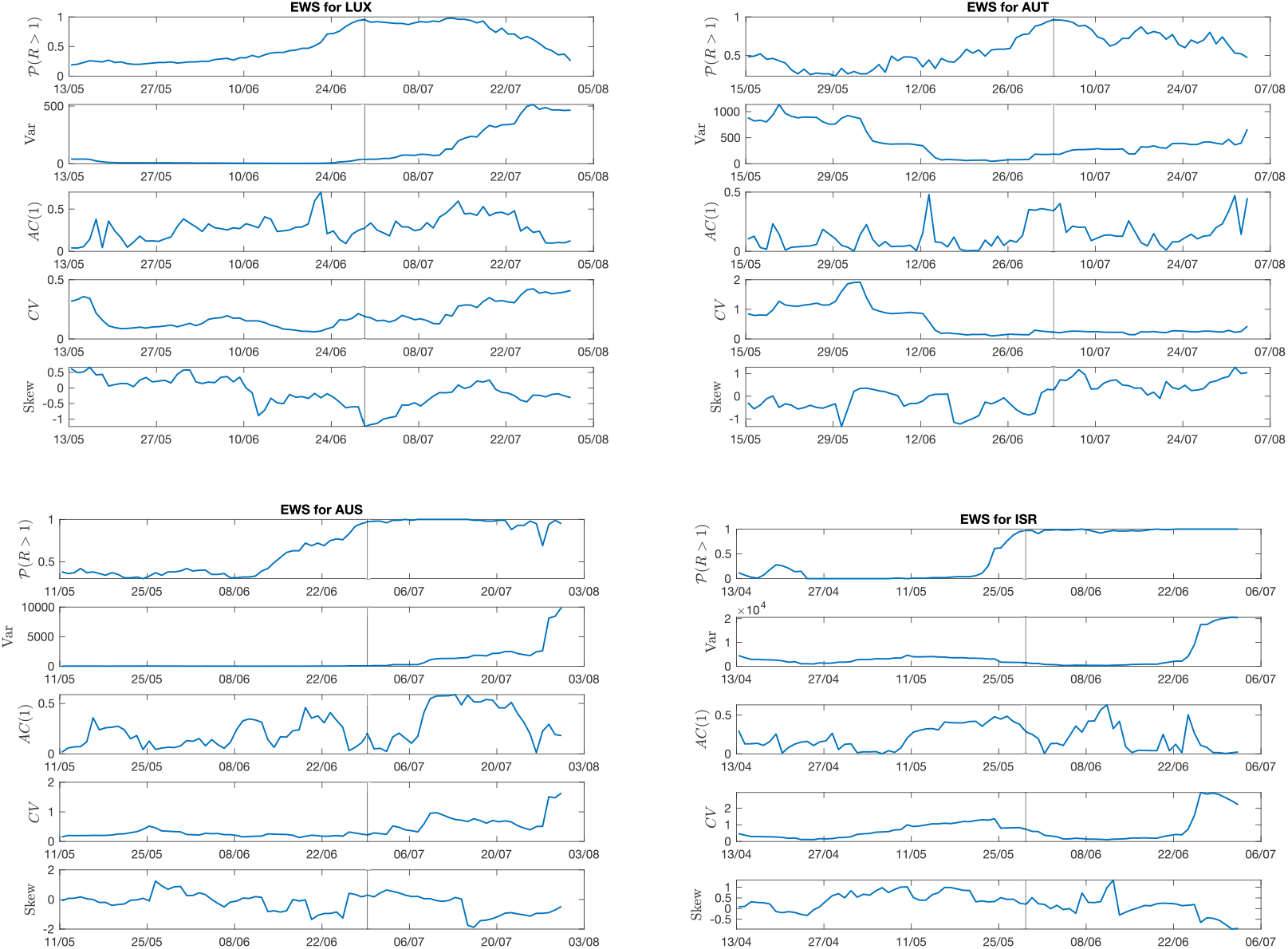
Evolution of EWS far from the transition point. Four example countries are shown: Luxembourg and Austria, with controlled features; State of Victoria (Australia), with small deviations from controlled features; and Israel that does not satisfy theoretical conditions. Considered EWS are the most common ones (variance, lag-1 autocorrelation, coefficient of variation, skewness). In addition, to mark the approach to the transition, *𝒫* (*R*(*t*) *>* 1) from the Bayesian estimation (see Eq. 5) is displayed. The vertical line reports the transition date. The detrending method here employed is Gaussian filtering. Other countries are reported in SI Appendix, as well as plots from ARIMA detrending.

For all indicators, the ARIMA detrending method yields better performances, quantified by higher AUC values. AC(1) is an exceptiion, as both methods return similar values, close to the ones of a random classifier. That ARIMA residuals yield higher AUC is likely due to the fact that ARIMA estimates the trends more closely at different time scales, thus returning more accurate fluctuations; on the contrary, the Gaussian filtering might be slightly more rough in considering only average time scales and returns approximated estimates for the fluctuations. This argument might explain why the skewness performs slightly better than a random classifier over ARIMA residuals: by considering more fine-grained time scales, the ARIMA seems able to pick the slight asymmetry in residual distributions that yield to skewness-related signals [56]. This analysis thus indicates the importance of choosing the detrending method to increase the detection performance of various indicators.

The same analysis was performed over the set *𝒩* of countries that do not satisfy the theoretical assumptions. Their AUC values are reported in Tab. 2. Such values clearly show that the considered indicators are not able to detect the transitions, overall performing worse than random classifiers. This supports what was already noticed for Israel in Fig. 3, where disrupted trends were observed, contradicting what was expected and thus returning a false negative signal. Time series for indicators of other countries in *𝒩* are reported in electronic supporting information Fig. S5. Hence, if a system is not known or there is difficulty in determining the type of data, incorrect conclusions could be drawn when interpreting the time series trend.

## 4 Discussion

Research on early warning signals from the theory of dynamical systems has greatly progressed in the last years, with a particular focus on disease re-emergence. However, verifying and interpreting empirical analysis according to theoretical assumptions has been so far limited. In this study, we observe that EWS from the critical transitions framework are able to detect the transition to disease re-emergence if basic theoretical assumptions like normal distribution of data fluctuations and slow change rates are satisfied. On the contrary, we confirm (along with [18]) that noise and commensurable time scales can obscure the early warning signals, which calls for caution when interpreting monitoring outputs. Hence, we suggest that statistical EWS can be suitable candidates for epidemic monitoring. Theory-based indicators can provide useful evidence, particularly when scarce data and few prior information are a constraint for using large scale statistics or Machine Learning (ML) methods. However, their alternate performance on unknown dynamics asks for careful assessment of the underlying dynamics. EWS have the potential to complement the existing toolbox of indicators to improve epidemic risk assessment but deserve further attention by scholars and decision-makers.

To support the growing corpus of theoretical studies, this study tested whether observed epidemic outbreaks behave consistently with the theory. When randomised experimental studies are not possible, observational studies provide stronger evidence if consistent patterns are seen in multiple locations and at multiple times, after checking for possible confounders. Hence, we employed world-wide available data about the ongoing COVID-19 pandemic, concentrating on the re-emergence of the disease after a first wave in Spring 2020. To limit biases associated with country-specific testing and reporting capacities, we constructed a limited, but curated set of time series data. Like *R*(*t*) and other indicators [57], EWS are estimates that rest on assumptions; hence, we screened the dataset to assess the matching of empirical features and theoretical assumptions. This pre-analysis showed that the same disease in diverse communities might have different noise distributions and evolve at different rates, which could depend on several factors including human behavior and population-wide interventions [58].

Furthermore, we tested how the best performing EWS detected the epidemic re-emergence. We carried out an extensive analysis of the effect of different detrending methods, showing that they are overall robust in highlighting the local trends of EWS. Such trends were studied both qualitatively and quantitatively, with ROC/AUC values. In particular, the ROC analysis assesses the robustness of online detection in distinguishing between real increasing trends and spurious fluctuations, which are often not studied in retrospective observations of time series data. In controlled settings, our results confirmed the expected trends in early warning signals and the potentials of the indicator system proposed. In particular, we showed that dynamical EWS are likely to operate successfully in contexts where the approach to the transition is gradual and not subject to high fluctuations. Further studies could associate these features to social behaviours and political strategies.

We also studied the different impact of detrending methods on the performance of warning signals. We showed that detrending time series with ARIMA models, appropriately calibrated for each country, increases the AUC score. This observation supports early theoretical works [45] and informs researchers about the importance of data processing methods to improve the performance of various indicators.

Finally, we analysed the potential limitations of the indicator system in other contexts, characterised by different dynamical features such as rapid increases of *R*(*t*) and strong or non-additive noise. This emphasises that, for EWS to properly work, the real system must fulfill the discussed conditions underlying the theoretical modelling. These open problems highlight that knowledge of the type of collected data is imperative to avoid misleading judgements in response to time series trends: EWS as epidemiological constructs will only remain valuable and relevant when used and interpreted correctly [59].

We acknowledge the limitations of this study, which might be overcome when new and better curated datasets will become available. First, data quality could be a limiting factor, despite being representative of real world monitoring capacities. In fact, the dataset selection highlights the importance of monitoring and of high quality prevalence data (as already suggested in [14]). Second, our definition of “ground truth” transition date is somewhat conservative, as we requested to have maximum probability of *R*(*t*), the control parameter, to be greater than 1. In the real world, the appropriate detection threshold is conditional on the various costs of a late outbreak alert, and requires an assessment by public health authorities which could modify the estimated lead time. Third, due to statistical uncertainties, a reliable estimation of the lead time - how much in advance a re-emergence can be predicted - was not entirely possible. Future studies will likely concentrate on this aspect, as early prediction would advance the current on-time detection.

In recognition that real epidemics might behave differently that what is commonly modelled, we nonetheless conclude that minimal dynamical models have the potential to predict relevant aspects of complex epidemics. While more detailed and complete multivariate models are being developed, macro-scale models based on complex systems theory can provide insights and indicators to detect epidemic re-emergence. On one hand, our results begin supporting the theoretical literature findings and their basic assumptions; on the other, they warn against naive applications of summary statistics as EWS: if not correctly applied, they could return possibly misleading spurious signals. In addition, our findings call for future studies on forecasting techniques based on pattern recognition in different dynamical regimes. For instance, validated EWS could serve as basis for the feature selection of automated Machine Learning-based algorithms [17]. The dual synergy of theoretical predictions and empirical studies will continue to play a role in the field of epidemic control and will likely have a impact in informing public health decisions.

## Data Availability

The analysis was performed in \textsc{Matlab} and Python. They require the \textit{Statistics Toolbox} and the \textit{scipy} library, respectively. Code and curated data are accessible on: \\ \url{https://github.com/daniele-proverbio/EWS_epidemic}.

https://github.com/daniele-proverbio/EWS_epidemic

## Author contribution

DP conceived and designed the study, collected the data, performed the analysis and interpreted the results. SM and FK performed the analysis and interpreted the results. JG supervised the project and interpreted the results. All authors wrote and approved the manuscript.

## Acknowledgments

The authors thank the Research Luxembourg - COVID-19 Taskforce for mutual collaborations, and Professors P. Ashwin and A. Skupin for their useful feedback. DP’s and SM’s work is supported by the FNR PRIDE DTU CriTiCS, ref 10907093. FK’s work is supported by the Luxembourg National Research Fund PRIDE17/12244779/PARK-QC. JG is partly supported by the 111 Project on Computational Intelligence and Intelligent Control, ref B18024.

## Code availability

The analysis was performed in Matlab and Python. They require the *Statistics Toolbox* and the *scipy* library, respectively. Code and curated data are accessible on: https://github.com/daniele-proverbio/EWS_epidemic.

## Supplemental Material for

### S1 Mathematical models and assumptions

As described in the Main Text, early warning signals (EWS) are measures that rest upon modelling assumptions. To clarify the most relevant assumptions, which are tested in the present study, we recall how EWS are theoretically derived.

When consistent with a mean-field approximation, the dynamics of COVID-19 infectiousness is well described by SIR-like models [1, 2]. To illustrate how early warning systems can be subsequently derived, we here recall the process described in [3]. SIR models describe disease processes in homogeneous populations of susceptible individuals (*X*), which can get infectious (*Y*) and eventually removed (*Z*) by death or recovery. In the deterministic SIR, transitions among states are governed by the infection rate *β* and the removing rate *γ*, which lumps recovery and death rate. Empirical values for COVID-19 can be traced in the abundant literature, e.g. [4]. In addition, systems are often open with influx rate *µ*^*1*^ and outflux rate *µ*^*″*^ of people. However, as many travelling restrictions^1^ were in place during the year 2020, we can assume that such fluxes are small and balanced: *µ*′ = *µ*″ = *µ*. Along with that, we model an influx rate of new cases *η* that can trigger subsequent disease outbreaks. Finally, intervention measures introduce a probability *p* that some susceptible individuals are isolated and protected, either physically or by vaccination [5]. The model reads:

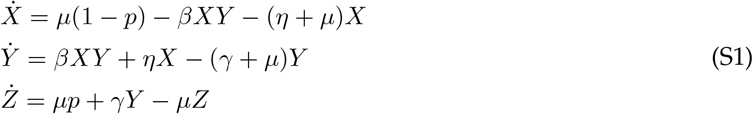

It is assumed that the population size *N* is constant, that is *X* + *Y* + *Z* = 1. In this case, the control parameter *R* is given by [3]:

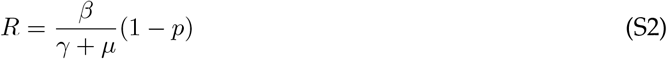

and reaches its critical value 1 for *p*^*\**^ = 1 − (*γ* + *µ*)*/β*, at which the dynamics undergo a transcritical bifurcation on the (*Y, p*) bifurcation diagram [3]. We remark that we consider here an example from literature: *R* might change in time after being driven by other evolving parameters such as one that tunes the contact rate *β* [2]. Without protection and extra fluxes, the basic reproduction number for COVID-19 was estimated at the beginning of the pandemic in the range 2 *< R <* 4 (*cf*., *e*.*g*., [6]).

If *p*(*t*) changes slowly over time, we can mathematically express the SIR model S1 approaching the transition as a slow-fast system:

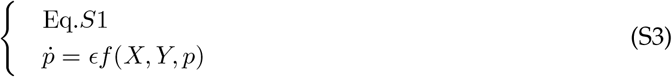

where 0 *< ϵ* « 1 and *f* is a function that describes its change. A limit case is ϵ → 0. This condition is necessary to interpret the dynamical shift as a slow crossing through a bifurcation point, and to compute its associated summary statistics [7]. Often [8, 3], it is assumed a constant 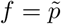. This way, the protection probability is, at first approximation, a linear function of time:

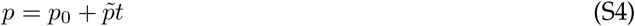

and *R* as well, following Eq. S2. If 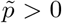, then *R* gets reduced and, if it was above 1, the bifurcation is crossed from above towards elimination. If 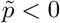,*R* increases and, if it was below 1, the bifurcation is crossed from below, towards a new emergence. Only the second case can be investigated with COVID-19 data, as most countries implemented suppression measures very rapidly [9] and thus Eq. S3 is not satisfied in the first case.

When the transition is approached from below, and if there are few cases, stochastic fluctuations are not negligible. Hence, we need to consider the transitions described by a stochastic master equation. Reducing it to Eq. S3 and a Fokker-Plank equation for the fluctuations was already performed in [3, 10]. Hence, we briefly recall them to illustrate the assumptions underlying the behavior of early warning signals prior to the transition.

First, note that system S1, along with condition 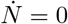, can be reduced to its first two equations. Hence, we just need to consider the transitions in and out *X* and *Y*. Second, for each small time step *dt*, the quasi-steady state *p* is constant.

Transitions in and out states are described as random jump processes. Such states are (*X, Y*), (*X* − 1, *Y*), (*X, Y* +1) and so on. Using *α* = (*X, Y*) to describe the “basic” state, 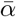 is any other state and *P* (*X, Y, t*) = Prob(*X*(*t*), *Y* (*t*) = (*x, y*)) is the probability that the state vector is equal to some non negative integer number (*x, y*). Finally, 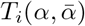 is the transition probability between states, and is a function of transition rates. The subscript *i* denotes all possible jumps in and out of the states. Examples of 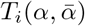 are found in [3, 11], depending on the system of interest. Consequently, the master equation for the stochastic process is:

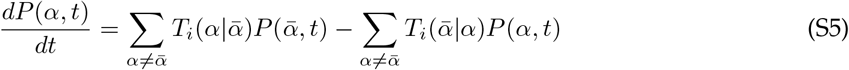

In general, Eq. S5 is nonlinear. To have analytical results about its average behavior and the fluctuations around it, the van Kampen expansion can be used [12] to approximate the discrete random variables with continuous random variables. This depends on having large *N*, which holds in our case when we consider the population of medium to big countries. The approximation of Eq. S5 is a system of normal random variables. To leading order, the expansion is equivalent to Eq. S3. To next-to-leading order, the obtained Fokker-Plank equation is equivalent to the following system of stochastic differential equations [10]:

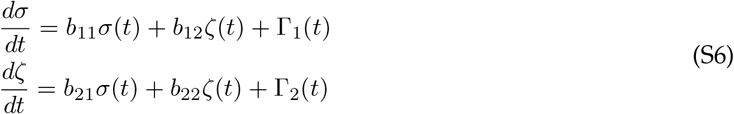

where *σ* and *ζ* are continuous random variables from the van Kampen expansion which represents the fluctuations from the susceptible and infectious states, respectively. Γ_*j*_ are white noise processes and the elements of the matrix *B* = {*b*_*kl*_*}* are functions of transition rates. Eq. S6 connects the stochastic description of the epidemic with SIR-like models like Eq. S1 with a noise term.

Eq. S6 can be analysed by its Fourier transform. By considering the fluctuations around the infectious state, we can derive the power spectrum and, through integration, the variance, the autocorrelation and other statistical moments. Their specific values depend on the eigenvalues of matrix *B* and of the covariance matrices of Γ_*j*_. The evolution of variance and autocorrelation next to the transition, as obtained in [3], is shown in Fig. S1 for a slow and a fast approach to *R* = 1. The trend of these summary statistic indicators on the fluctuations, prior to the transition, constitutes the set of signals that could detect the transition itself; for instance, the increase in variance, often measured in terms of Kendall’s *τ*. The Kendall’s *τ* score is a non-parametric measure of ranks correlation, which is usually used to identify increasing trends [13, 14]. Such increasing trends are known in the literature as early warning signals (EWS) [15].

Finally, let us recall the general theory of EWS on bifurcations. Any system approaching a transcritical bifurcation is, in its vicinity, topologically equivalent to a transcritical normal form [16, 17]. This is a minimal model that retains the systems’ behavior and resilience properties in the vicinity of a bifurcation. Models can be transformed to a normal form after an appropriate change of variables [18]. The one associated to a transcritical bifurcation has the form:

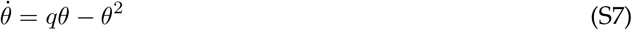

where *q* is the bifurcation parameter and *θ* the variable of interest (*Y* from Eq. S1, in this specific case). This form represents a system whose extinction state and positive steady state coalesce and exchange stability when *q* reaches its critical value. If there are statistical fluctuations *ξ*, we can write their evolution as a linearization around the equilibrium 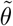, thus resulting in a Langevin equation [7, 19]:

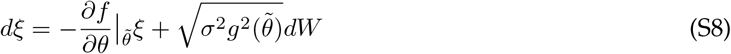

where *dW* is a Wiener process, *σ*^2^ models the noise level and *g* is the diffusion coefficient of the associated Fokker-Planck equation. With a linear 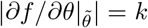, Eq. S8 is an Ornstein–Uhlenbeck process with known statistical moments [20]. For instance, the theoretical variance is:

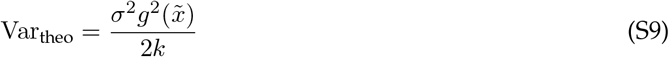

and the theoretical autocorrelation is:

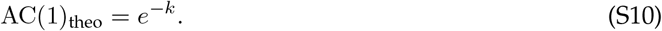

Hence, we can complement the theoretical results described above with those from the theory of statistical indicators of normal forms, e.g. [7, 19, 21]. Important remarks are that: a) multiplicative noise can modify the indicators trend, e.g. by making it decrease; b) EWS are expected to work best in the vicinity of the transition; c) there can be bifurcation delays associated to out-of-equilibrium phenomena, i.e. changes of the system state (and of its indicators) that lag behind the bifurcation of the limit case.

**Fig. S1.**
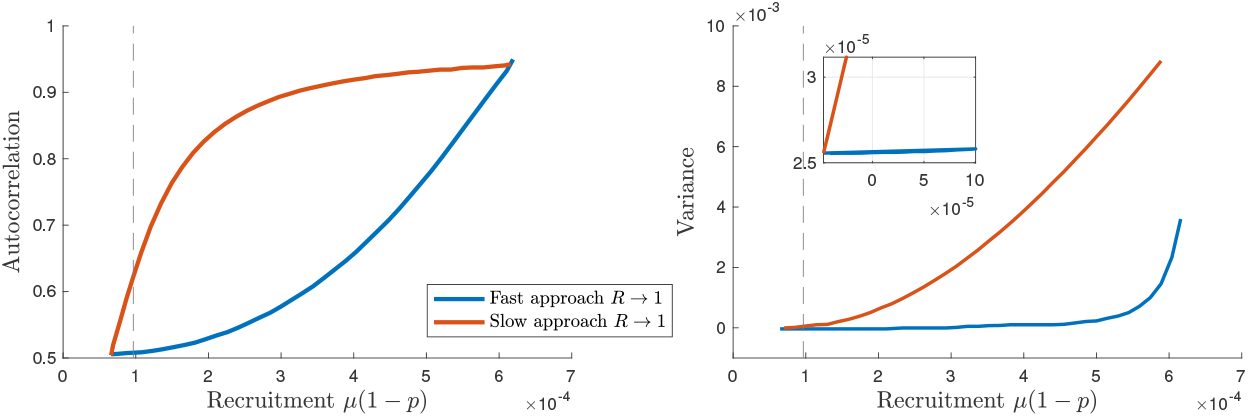
Theoretical EWS for epidemic re-emergence. Left: the lag-1 autocorrelation increases before the transition if the approach of *R* to 1 is slow (slow-fast assumption satisfied, red); otherwise it increases after the transition (bifurcation delay, blue). Right: the variance increases before the transition (dashed grey line) if the approach of *R* to 1 is slow (slow-fast assumption satisfied, red; the inset magnifies the effect). Otherwise, it increases after the transition (bifurcation delay, blue). The plots are derived from the analytical results of [3] and reproduce its Fig. 9, d−e. On the x-axis: values for the recruitment rate of new infectious that could trigger a re-emergence.

### S2 Data collection and curation: examples

Among all the countries that registered a re-emergence of COVID-19 epidemic between April and September 2020, we first selected those for which meaningful prevalence data could be directly obtained from official sources or reconstructed with Eq. 2 from Main Text. Examples of discarded data series are reported in Fig. S2. We recall the underlying hypothesis of this study: that dynamical early warning signals are expected to work when the investigated system can be described by a proper dynamical model. Hence, we did not consider time series for which active cases do not display the typical SEIR-like behavior like that described in [22, 23, 2] (mostly due to data management and reporting), or for which recovered cases are reported with different frequencies, resulting in sawtooth curves for active cases. In the later case, the detrended fluctuations would be associated to reporting standards and would not be representative of dynamical fluctuations associated to EWS. Other selecting criteria to increase the quality of the dataset are discussed in “Materials and Methods” of the Main Text.

Curves of active cases for all countries considered in Main Text, their smoothing and the associated *R*(*t*) are displayed in Fig. S3. The smoothed curves serve as basis for the detrending, the analysis of the noise distribution of each country and the subsequent investigation of early warning signals, as explained in Main Text.

**Fig. S2.**
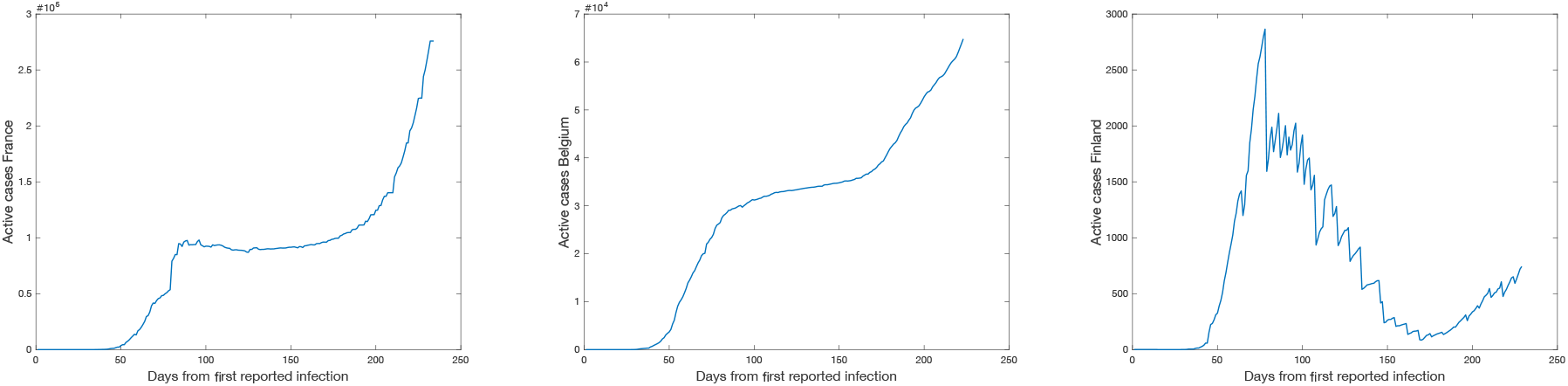
Examples of discarded time series, following some of the criteria explained in “Material and Methods” of Main Text. Among others, France and Belgium had active cases curves that differ from the typical bell-shaped SEIR-like behavior. This is related to reporting of recovered and dead patients. On the other hand, Finland is an example of sawtooth evolution, due to recovered cases being reported with different frequencies than daily cases. Data from [24].

### S3 Estimating R(t) with Bayesian MCMC

Following standard methodologies [25, 26], we reconstruct the day-by-day evolution of the reproduction number *R*(*t*) by fitting a Poisson transmission model with Markov Chain Monte Carlo (MCMC) methods.

When modelling “arrivals” of discrete-state stochastic processes (*cf*. section S1), Poisson processes are widely employed. For instance, it effectively modeled the transmission of Ebola [27] and Influenza [28]. It states that, given an average rate of *λ* new cases per day, the probability of seeing *k* new cases is distributed according to the Poisson distribution:

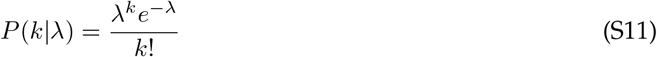

In turn, *λ* depends on *R* as [29]:

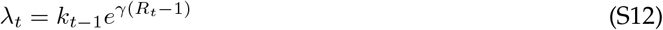

where *γ* is the reciprocal of the serial interval, which is around 4 days for COVID-19 [30, 31]. To account for such uncertainty, we treat *γ* as a random sample from a Gaussian distribution centered in 4 days with standard deviation 0.2. Hence, the probability of observing a time series *x* = {*x*_*t*_*}* between *t*_0_ and *T* is given by:

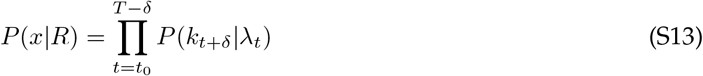

for each time step *δ*. Before *t*_0_, no cases were reported. Following Bayes’ rule, the posterior distribution of *R*, for each time point, is given by (up to a normalization constant):

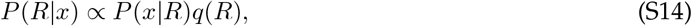

where *q*(*R*) is a prior distribution. For each time point after *t*_0_ +1, the prior equals the preceding posterior. We follow he implementation of [32] to generate thousands of MCMC samples with Metropolis Hastings, starting with a Gaussian prior *𝒩* (*R, σ*). We used *σ* = 0.15 as it maximizes the log-likelihood over every state in the current implementation. From the posterior distribution, we also estimated the probability that *R*(*t*) is greater than 1, which was in turn used to define the “ground truth” date of regime transition for the epidemic trend (see also “Materials and Methods” of Main Text for further discussion). Fig. S3 shows the results of the Bayesian *R*(*t*) estimation (median values and 50% Credible Intervals) for the considered countries.

### S4 Determining the rate of approach for R(t) → 1

As explained in the Main Text, we are interested in evaluating how rapidly the transition point is approached, for each country. To do so, the simplest linear trend

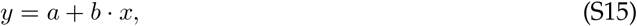

corresponding to Eq. 13 in Main Text, is assumed and fitted to *R*(*t*) time series, obtained as described above and reported in Fig. S3. The estimated regression coefficient is informative about the rate of approach of *R*(*t*) → 1.

The fitting was performed with *scipy*.*optimize* routine, considering as uncertainty the 50% credible interval from the distribution of *R*(*t*), which is representative of one standard deviation. The goodness of fit was evaluated with the reduced *χ*^2^ score. Results are displayed in Fig. S4. The 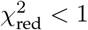 guarantees the goodness of the linear fit, which allows to extrapolate the regression coefficient as a measure of the rate of *R*(*t*) → 1. The values of *b* ± *σ*_*b*_ are then reported in Fig. 1c of Main Text.

### S5 Evolution of EWS for all countries

In this section, we show and discuss the evolution of the considered early warning indicators for all countries, including those that are not shown in Fig. 3 of Main Text.

Let us first consider EWS obtained from Gaussian filter detrending. In Fig. S5 we observe the evolution of the indicators, either globally (S5a) or locally, just prior to the bifurcation (S5b for the variance, the most robust one as discussed in Main Text). We can observe the patterns discussed in the Main Text, associated to the different countries belonging to the test set *𝒴* or not (*𝒩*). Within *𝒴*, the variance increases prior to the transition and gives very few spurious signals before, whereas other indicators can be more misleading when the transition still did not happen. Overall, predicted trends of EWS prior to the bifurcation are associated to satisfying theoretical assumptions such as gradual approach of *R*(*t*) → 1 and white noise (*cf*. Fig. 1 in Main Text and discussion thereafter). Not satisfying these requirements might disrupt the expected increasing trend and results in misleading signals, see in Fig. S5 the countries listed in. *𝒩* Hence, if a system is not known or there is difficulty in determining the type of data, incorrect conclusions could be drawn when interpreting the time series trend. Fig. 4 and Table 2 of Main Text then quantify the performance of EWS for all countries.

**Table 2:** AUC scores for different indicators, over 𝒴 and 𝒩 datasets, after Gaussian or ARIMA detrending methods.

| Indicator | Gaussian det. |  | ARIMA det. |  |
| --- | --- | --- | --- | --- |
| | over $\mathcal{Y}$ | over $\mathcal{N}$ | over $\mathcal{Y}$ | over $\mathcal{N}$ |
| Variance | 0.6671 | 0.1981 | 0.7468 | 0 |
| AC(1) | 0.5258 | 0.4995 | 0.5182 | 0.3063 |
| CV | 0.3968 | 0.1043 | 0.4375 | 0 |
| Skewness | 0.4664 | 0.2925 | 0.5955 | 0.4974 |

### S6 ARIMA detrending and corresponding global EWS

As explained in the Main Text, on top of moving average smoothing and Gaussian kernel filtering, we tested the ARIMA detrending method. ARIMA (Autoregressive Integrated Moving Average) is an automatic method to identify the leading trends of a time series [33], depending on three terms (*p, q, d*) that need to be adjusted for each data set. Initially, the *d* term is identified by checking for stationarity in the data. Then, the other terms are automatically identified with the python function *pmdarima* for ARIMA estimation.

For State of Victoria (Australia), we use ARIMA(0,2,1) whose residuals have mean -2.51 and standard deviation 197.9. For Austria, we use ARIMA(1,2,0) whose residuals have mean 2.58 and standard deviation 121.5. For Demark, we use ARIMA(0,2,1) whose residuals have mean 3.17 and standard deviation 86.97. For Israel, we use ARIMA(3,2,2) whose residuals have mean 34.21 and standard deviation 823.5. For Japan, we use ARIMA(1,2,2) whose residuals have mean -0.2788 and standard deviation 349.12. For Korea, we use ARIMA(3,1,3) whose residuals have mean 6.462 and standard deviation 141.14. For Luxembourg, we use ARIMA(2,1,3) whose residuals have mean 0.3402 and standard deviation 30.90. For Nepal, we use ARIMA(1,2,0) whose residuals have mean 0.248 and standard deviation 241.66. For Singapore, we use ARIMA(1,2,0) whose residuals have mean -0.0698 and standard deviation 152.47. For Veneto, we use ARIMA(0,2,1) whose residuals have mean 0.1330 and standard deviation 70.048. The residual of state of Victoria, Austria, Denmark, Israel, Japan, Luxembourg, Nepal, Singapore and Veneto correspond to white noise as the portmanteau test returns a large p-value.

Fig. S6 displays the plots for global behavior of statistical indicators obtained after ARIMA detrending. It corresponds to Fig. 4 of main text. We observe that the trends are similar across the two figures, but slightly more stable on the ARIMA side, as quantified by the ROC curve Fig. 5b in Main Text.

**Fig. 5.**
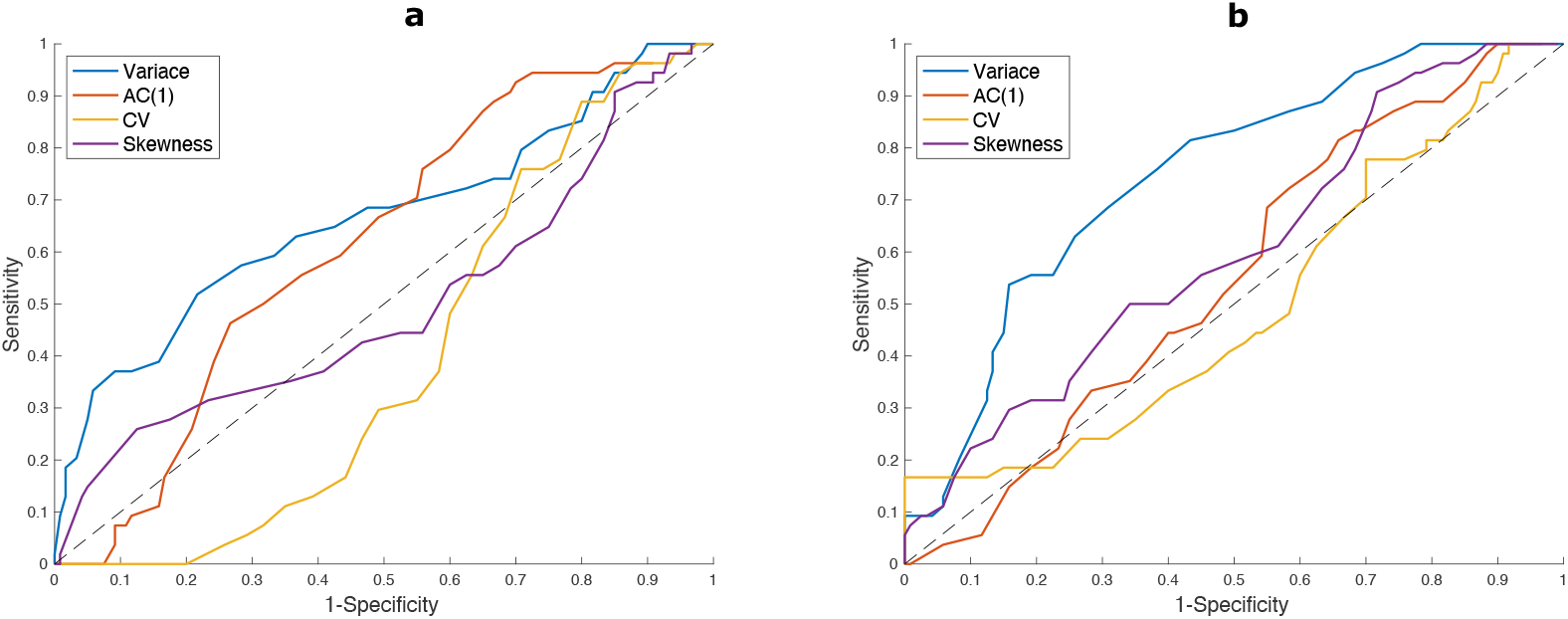
ROC curves for each considered indicator, estimated on the test set. Each point corresponds to a test value for *τ*, to define if the detection if positive. The diagonal line corresponds to the ROC of a random classifier. Curves above it imply better performance. As explained in Materials and Methods, the curves represent an average over the countries included in the test set. a) Computed on Gaussian filtered data; b) Computed on ARIMA detrended data.

**Fig. S3.**
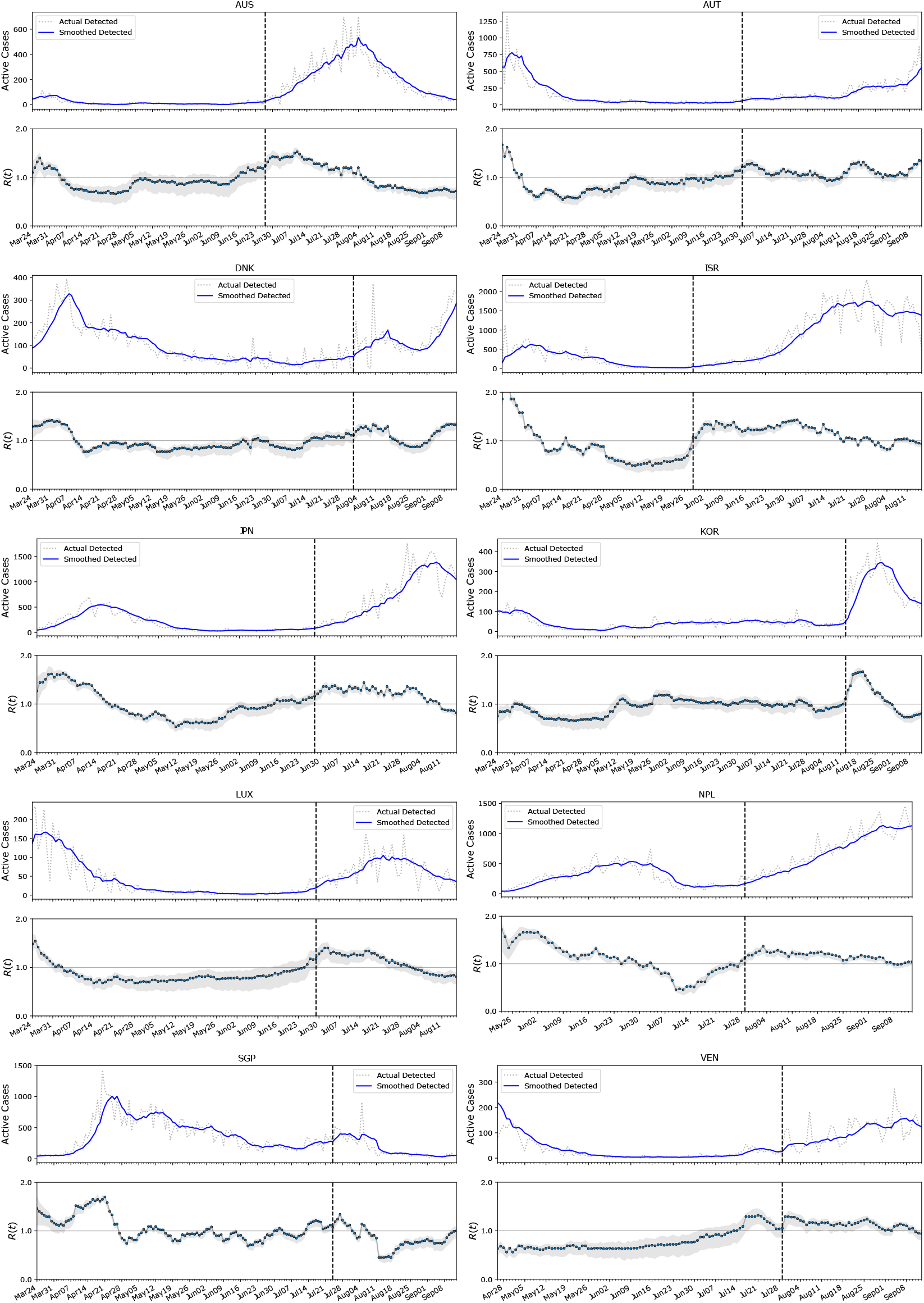
Curves of active cases for the considered countries, along with their associated *R*(*t*) (median values and 50% Credible Intervals). The vertical dashed line identifies the day marked for the transition an reported in Table 1 of Main Text.

**Fig. S4.**
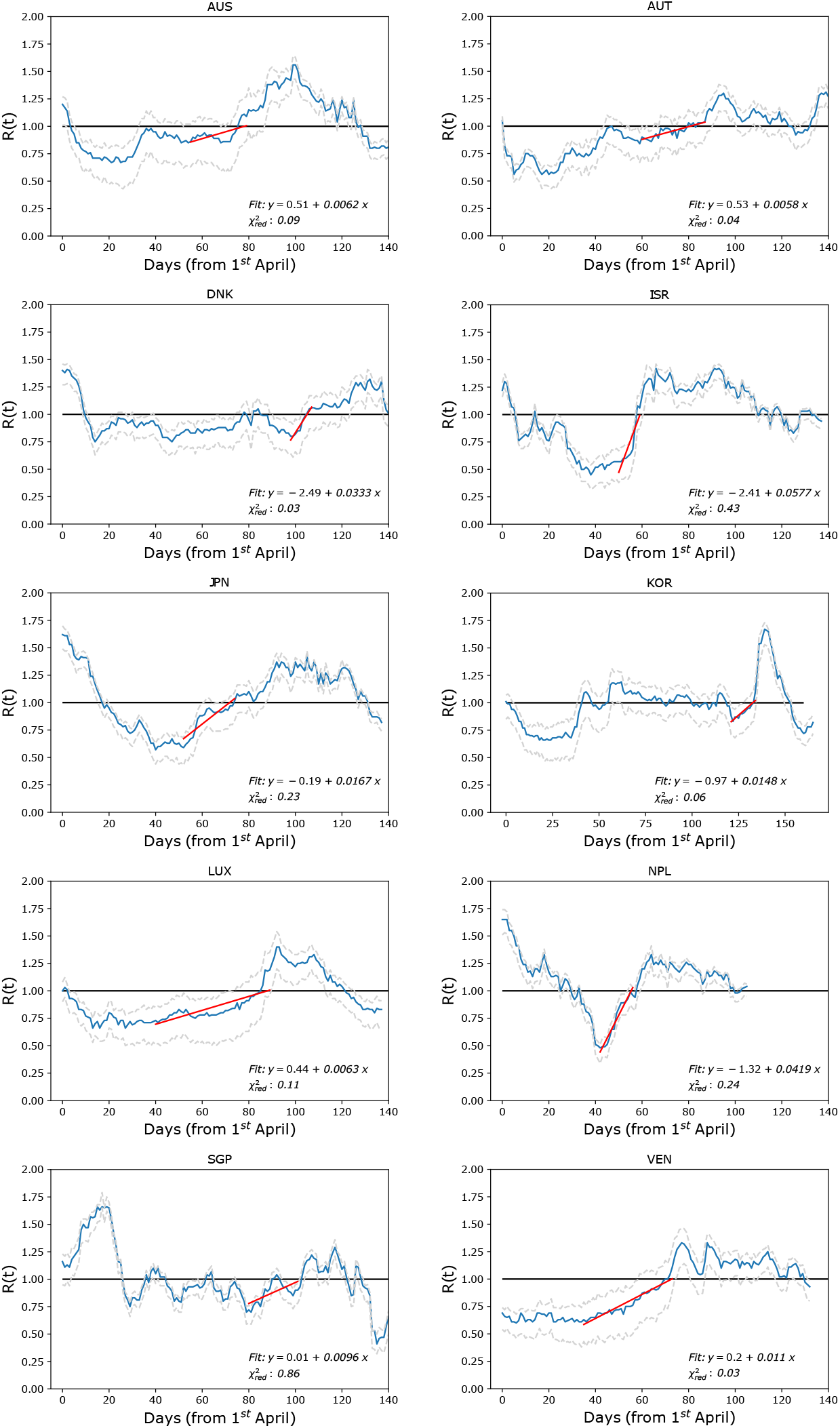
Fitting *R*(*t*) evolution (blue line; dashed lines are ±50% CI) with a linear trend prior to the transition, to estimate the rate of approach to the threshold value 1. The fit begins around the minimum of *R*(*t*) (excluding small fluctuations) until when the median value crosses 1 (horizontal line). The best fit curve is in red. 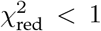 guarantees that a good fit is achieved for the simple linear function Eq. S15. The regression coefficient *b* is a proxy for the rate of approach to 1. Its associated uncertainty is not reported here but is shown in Fig. 1 of Main Text.

**Fig. S5.**
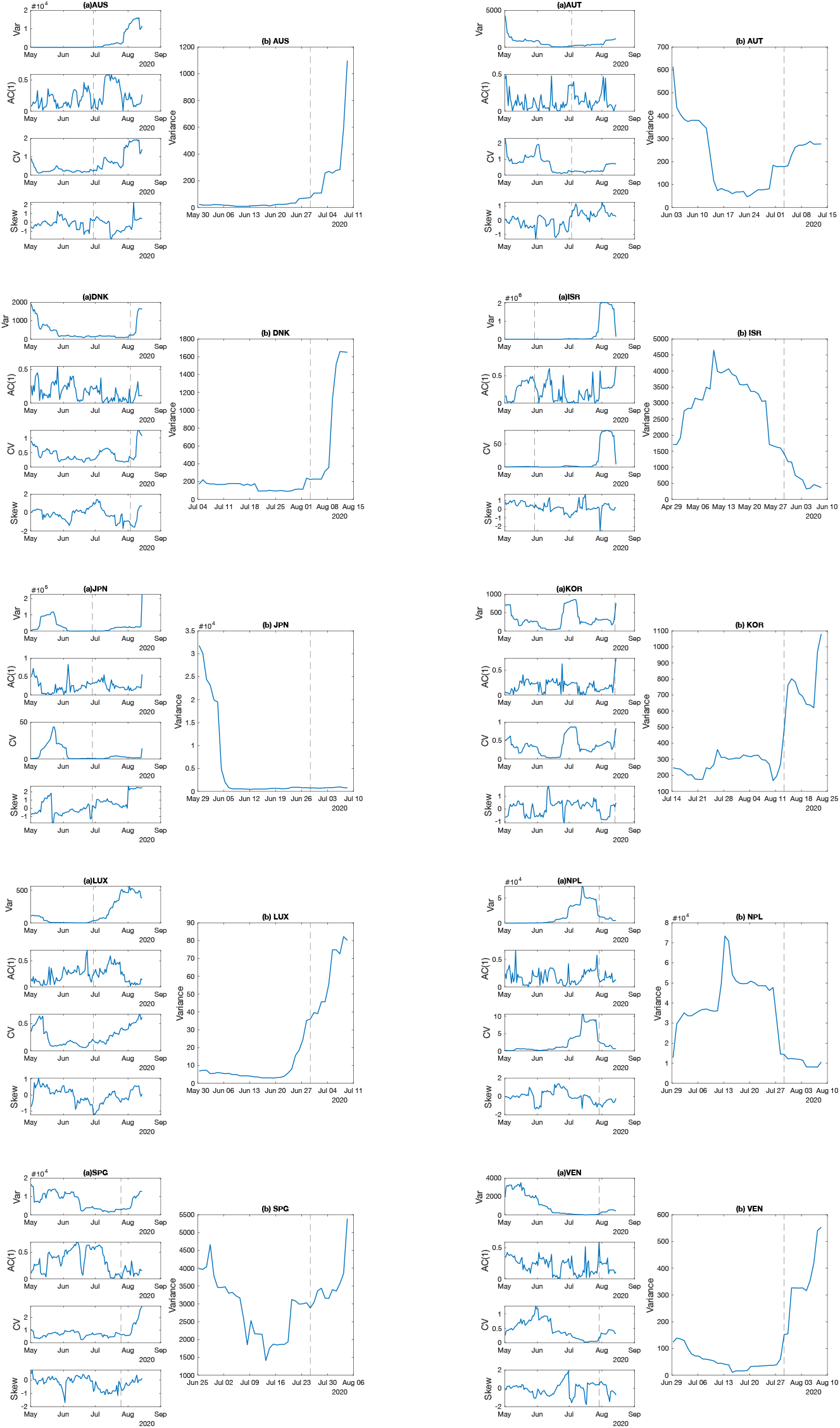
Evolution of the considered indicators for all countries. Figs (a) report their global evolution, from the end of the first wave to the second. Figs (b) focus on the behavior of the variance close to the transition (local behavior). As discussed in the Main Text, we report the values of indicators after a moving window of size 14 days, associated to the rightmost data point to avoid “looking into the future”. The vertical dashed line marks the transition point.

**Fig. S6.**
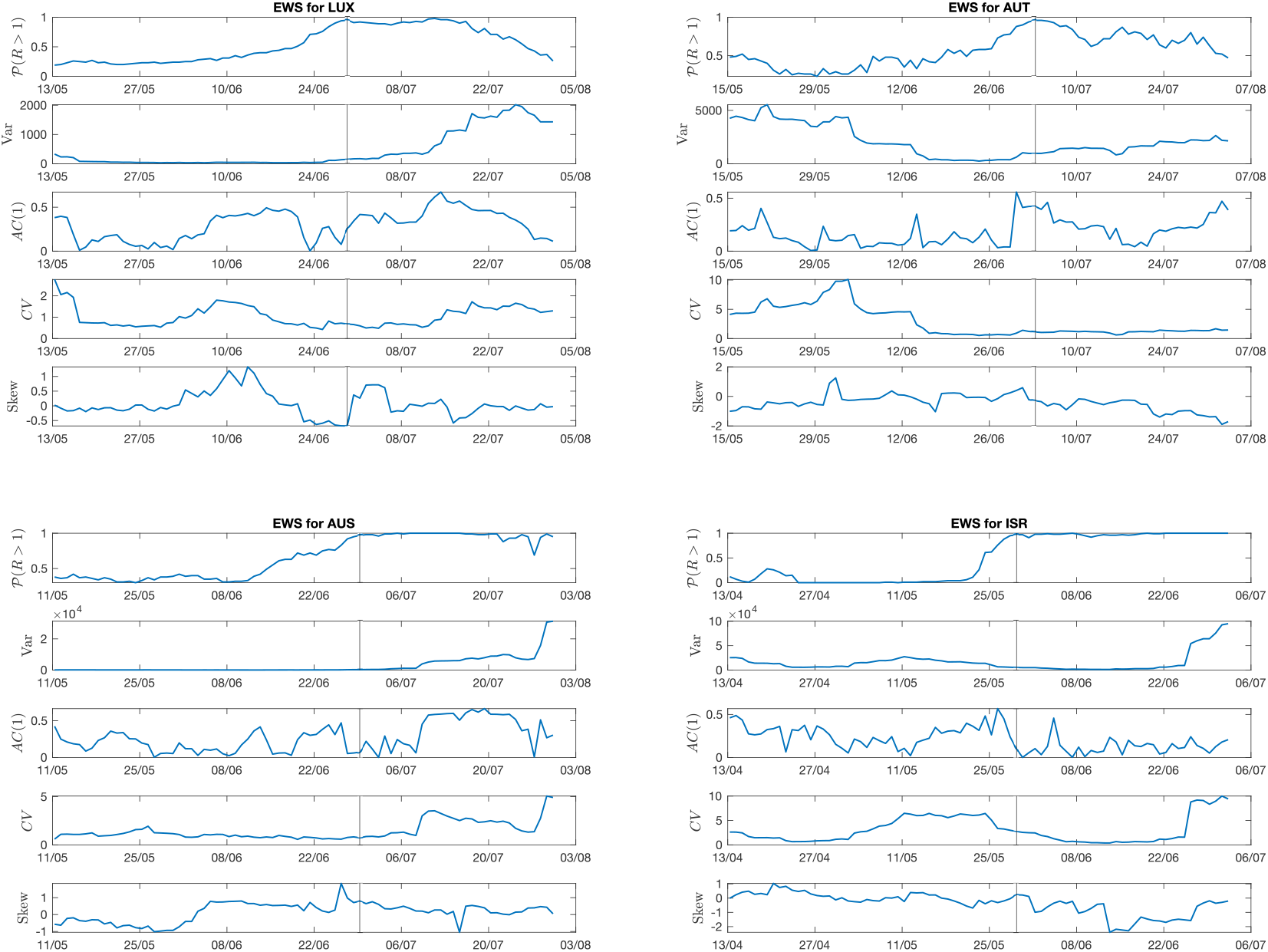
Evolution of EWS far from the transition point. Four example countries are shown: Luxembourg and Austria, with controlled features; State of Victoria (Australia), with small deviations from controlled features; and Israel that does not satisfy theoretical conditions. Considered EWS are the most common ones (variance, lag-1 autocorrelation, coefficient of variation, skewness). In addition, to mark the approach to the transition, 𝒫(*R*(*t*) *>* 1) from the Bayesian estimation (see Eq. **??**) is displayed. The vertical line reports the transition date. The detrending method here employed is ARIMA.

## Footnotes

1 Refer to the ACAPS website (https://bit.ly/3nFFqUS) for a dataset of government measures.

